# Benchmarking Machine Learning Missing Data Imputation Methods in Large-Scale Mental Health Survey Databases

**DOI:** 10.1101/2024.05.13.24307231

**Authors:** Preethi Prakash, Kelly Street, Shrikanth Narayanan, Bridget A. Fernandez, Yufeng Shen, Chang Shu

**Affiliations:** Department of Computer Science, Columbia University, New York, NY, USA; Division of Biostatistics, Department of Population and Public Health Sciences, Keck School of Medicine, University of Southern California, Los Angeles, CA, USA; Viterbi School of Engineering, University of Southern California, Los Angeles, CA, USA; Division of Medical Genetics, Department of Pediatrics, Children’s Hospital Los Angeles and The Saban Research Institute, Los Angeles, CA, USA; Department of Pediatrics, Keck School of Medicine of USC, University of Southern California, Los Angeles, CA, USA; Department of Systems Biology, Department of Biomedical Informatics, and JP Sulzberger Columbia Genome Center, Columbia University Irving Medical Center, New York, NY, USA; Center for Genetic Epidemiology, Division of Epidemiology and Genetics, Department of Population and Public Health Sciences, Keck School of Medicine, University of Southern California, Los Angeles, CA, USA

**Keywords:** Missing data, mental health survey, imputation, machine learning

## Abstract

Databases with mental and behavioral health surveys suffer from missingness when participants skip the entire survey, affecting the data quality and sample size. We investigated the missing data patterns and evaluate the imputation performance in Simons Powering Autism Research (SPARK), a large-scale autism cohort consists of over 117,000 participants. Four common methods were assessed – Multiple Imputation by Chained Equations (MICE), K-Nearest Neighbors (KNN), MissForest, and Multiple Imputation with Denoising Autoencoders (MIDAS). In a complete subset of 15,196 autism participants, we simulated three types of missingness patterns. We observed that MIDAS and KNN performed the best as the rate of random missingness increased and when blockwise missingness was simulated. The average computational times for MIDAS and KNN were 10 minutes, 35 minutes for MissForest, and 290 minutes for MICE. MIDAS and KNN both provide promising imputation performance in mental and behavioral health survey data that exhibit blockwise missingness patterns.

## Introduction

Large-scale biobank databases in mental and behavioral health such as Simons Powering Autism Research for Knowledge (SPARK), UK Biobank and All of Us have empowered researchers to investigate the genetic and environmental risk factors associated with mental and behavioral disorders among more than 100,000 subjects ^1-3^. Self-reported surveys and questionnaires such as the Social Communication Questionnaire (SCQ) ^4^, Repetitive Behavior Scale-Revised (RBS-R) ^5^ and Developmental Coordination Disorder Questionnaire (DCDQ) ^6^ are commonly used to quantify mental and behavioral functions at scale. These questionnaires typically consist of a series of related questions and measure responses using ordinal scales with a natural order or rank to indicate level of agreement known as Likert scales ^7^.

However, missingness commonly occurs in the responses to these surveys and questionnaires. The reasons include non-inapplicable or ambiguous questions, and characteristics of the participants themselves including reluctance to answer sensitive questions, incomplete knowledge, and lack of time. Missingness can also arise at the source level. Specifically, data may have been curated from varying sources with different administered instrument protocols. Certain questions in the survey also may not be relevant to specific demographic groups, such as those that might not apply to young children.

Common types of missing data include Missing Completely at Random (MCAR) and Missing Not at Random (MNAR) with either specific parts of surveys or entire surveys being incomplete ^8^. In MCAR, the probability of missingness is independent of the observed and unobserved data. MAR is a broader class than MCAR in which the missing data is related to the observed but not the unobserved data. On the other hand, the probability of missingness in MNAR data depends on the unobserved missing values. Typically, participants tend to skip entire questionnaires due to unobserved factors, and a form of MNAR missingness referred to as blockwise missingness arises. Blockwise missingness occurs when all responses belonging to the same survey are missing simultaneously for the same participants, forming clustered missing blocks in the overall phenotypic data.

The simplest solution to address blockwise missingness in mental and behavioral questionnaires is to drop participants with missing surveys ^9^. However, this option leads to a significant loss of information, reduced sample size and loss of statistical power when analyzing mental and behavioral questionnaires in biobank data. Another commonly used approach is to impute missing data using statistical and computational methods. Mean, median, and mode substitutions are basic imputation approaches that maintain the original sample size but can lead to biased inferences ^10^. Specifically, participants who skip certain questionnaires may exhibit different characteristics than those who complete the questionnaires ^11^.

More advanced imputation approaches using statistical and computational methods are needed to accurately impute mental and behavioral surveys with blockwise missingness. Here, we employed four commonly used missing data imputation methods - Multivariate Imputation by Chained Equations (MICE), K-Nearest Neighbors (KNN), nonparametric missing value imputation using Random Forest (MissForest), and Multiple Imputation with Denoising Autoencoders (MIDAS) ^12-15^. MICE is one of the most popular methods of multiple imputation originally developed in the early 2000s ^12^. This approach uses a series of regression models to predict each variable with missingness using the remaining variables in the data ^13^. KNN is a supervised machine learning algorithm commonly used when the distribution of the data is unknown or difficult to determine^14^. This method performs predictions on the missing data by averaging the *k* nearest data points. Nonparametric Missing Value Imputation using Random Forest (MissForest) is a missing data imputation method based on random forest developed in 2012. It predicts missing values based on random forest models trained on the complete dataset and imputes missing values iteratively ^15^. Multiple Imputation with Denoising Autoencoders (MIDAS) uses a type of unsupervised neural network to predict missing values in the data by reducing the dimensions in the observed data and reconstructing the missing data. MIDAS was recently developed in 2022 and has proven its high accuracy and computational efficiency through systematic tests on simulated and real social science data ^16^.

Previous studies have not systematically reviewed new imputation methods in the databases with mental and behavioral health surveys^17-21^. Additionally, they have not focused on assessing imputation accuracy in surveys with blockwise missing structures^17-21^. This study systematically examines the imputation performance and computational time of these four commonly used missing data imputation methods (MICE, KNN, MissForest and MIDAS) in the presence of blockwise missingness in mental and behavioral surveys. It uses data from the Simons Powering Autism Research for Knowledge (SPARK), a large-scale autism research study that collects social functioning and behavioral surveys from over 117,000 participants. This study assesses imputation models on both MCAR and MNAR data, identifying the optimal method for each type of missingness pattern. This study conducts a novel exploration of these methods while also addressing the commonly encountered blockwise missingness pattern.

## Methods

Figure 1 outlines the sample selection and workflow of the study. The four major steps included (1) preprocessing the data to generate a dataset comprised of complete observations, (2) setting up the simulation scenarios for three missing data mechanisms including random missingness, survey-specific missing rates, and blockwise missingness with survey-specific missing rates, (3) conducting the missing data imputation, and (4) evaluating the performance of each model.

**Figure 1.**
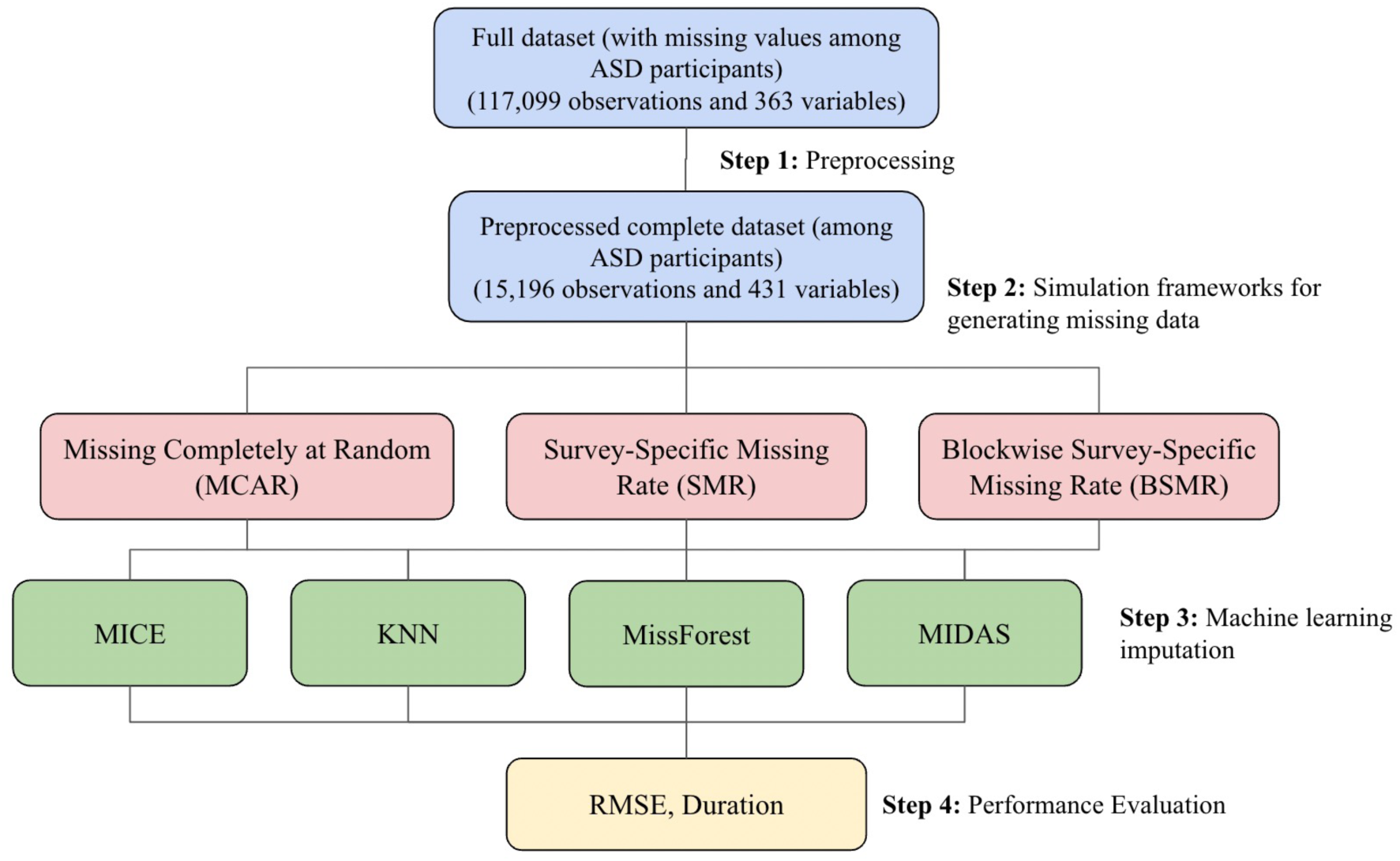
Overview of workflow and study design. a) The full dataset refers to the original data filtered to only include ASD participants. The preprocessed complete dataset refers to the original dataset after filtering to only include ASD participants, dropping incomplete rows, removing variables with extreme rates of missingness, and conducting one-hot-encoding on the categorical variables (which increases the number of variables). b) MCAR refers to the simulation scenario which randomly converts a specified fraction of the input dataset to missing. SMR refers to the simulation environment that is tailored to the missingness of the original dataset. BSMR refers to the simulation environment that is also tailored to the missingness of the original dataset, but converts all rows of a given column to missing at once. c) MICE is an imputation method that employs a series of regression models; MissForest is an imputation method that is based on random forests; MIDAS is an imputation method that uses denoising autoencoders; KNN is an imputation method that uses neighboring data points in the feature space. d) RMSE corresponds to Root Mean Squared Error.

### 1. Data Source and Preprocessing

The dataset used in this study is based on SPARK phenotype V8, with 117,099 participants with autism and 363 variables. It contains information extracted from standardized surveys and parent-reported medical history regarding children with autism. The following 8 surveys with <80% missing rates in the full dataset (**Table 1**) were included in missing data imputation assessment: Individuals Registration, Basic Medical Screening, Background History, Social Communication Questionnaire (SCQ), Repetitive Behavior Scale-Revised (RBS-R), Developmental Coordination Disorder Questionnaire (DCDQ), Child Behavior Checklist (CBCL), and Area Deprivation Index (ADI).

**Table 1.**
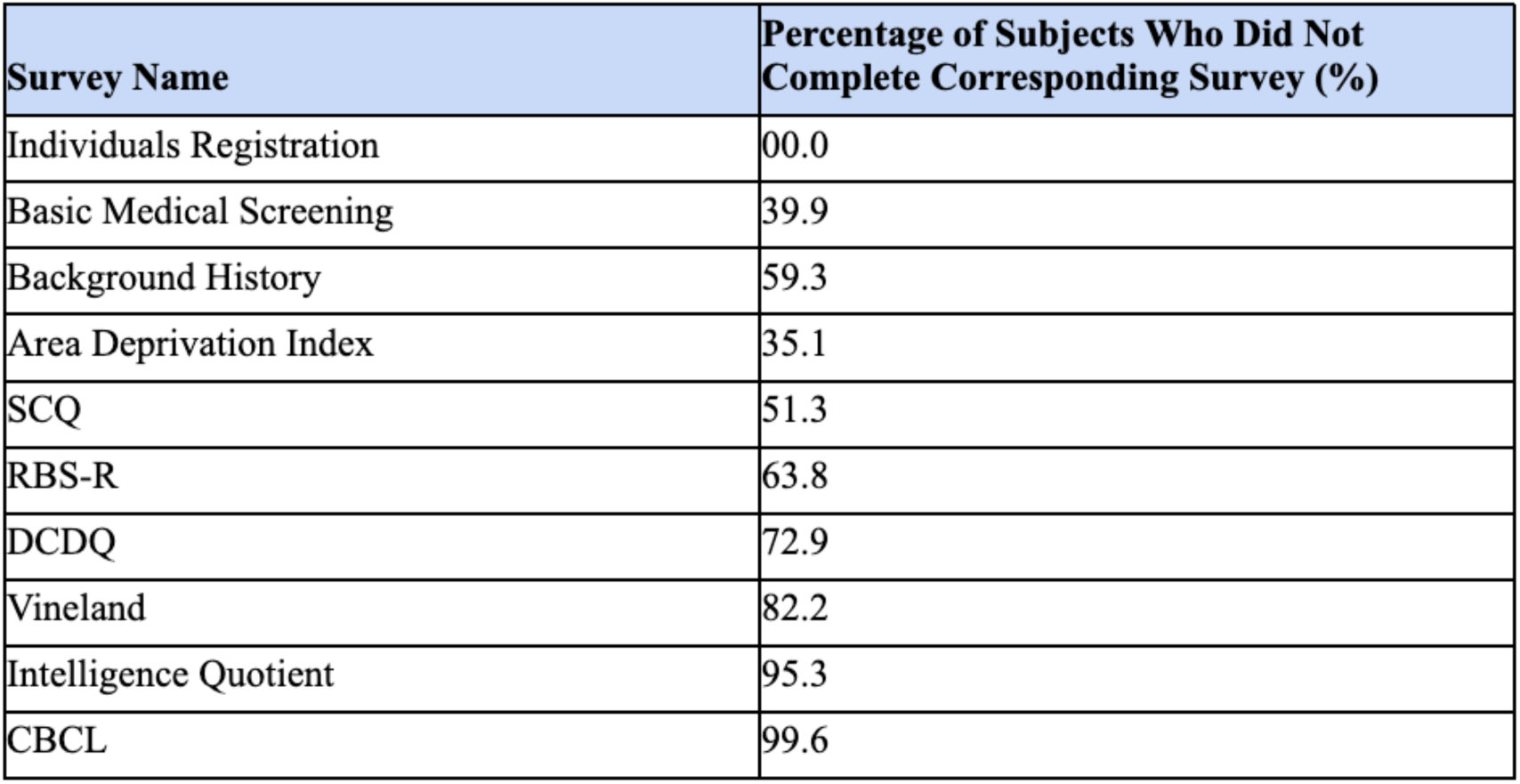
Percentage of subjects who did not complete each individual survey among all 117,099 participants with autism in SPARK. Social Communication Questionnaire (SCQ), Repetitive Behavior Scale-Revised (RBS-R), and Developmental Coordination Disorder Questionnaire (DCDQ) are surveys commonly used to quantify the mental and behavioral functions at scale.

| Survey Name | Percentage of Subjects Who Did Not Complete Corresponding Survey (%) |
| --- | --- |
| Individuals Registration | 00.0 |
| Basic Medical Screening | 39.9 |
| Background History | 59.3 |
| Area Deprivation Index | 35.1 |
| SCQ | 51.3 |
| RBS-R | 63.8 |
| DCDQ | 72.9 |
| Vineland | 82.2 |
| Intelligence Quotient | 95.3 |
| CBCL | 99.6 |

This dataset was first filtered to remove variables with extreme rates of missingness (∼90% or greater), resulting in a drop of 22 variables. The dataset was then modified to remove any rows with missing information. This resulted in 15,196 participants with autism and 347 variables.

One-hot encoding was used to transform the categorical variables in this dataset, resulting in 15,196 participants with autism and 431 variables. The `preProcess` method from the `caret` package in R was used to center and scale each column to have a mean of 0 and standard deviation of 1. This was mainly to allow for comparable Root Mean Squared Error (RMSE) metrics across all variables.

This preprocessed complete dataset of participants with autism was used to simulate different missing data mechanisms and assess the accuracy or various imputation methods.

### 2. Three Simulation Scenarios for Missing Data Mechanisms

We simulated three simulation scenarios for missing data mechanisms in mental and behavioral surveys as outlined below and in Figure 2.

**Figure 2.**
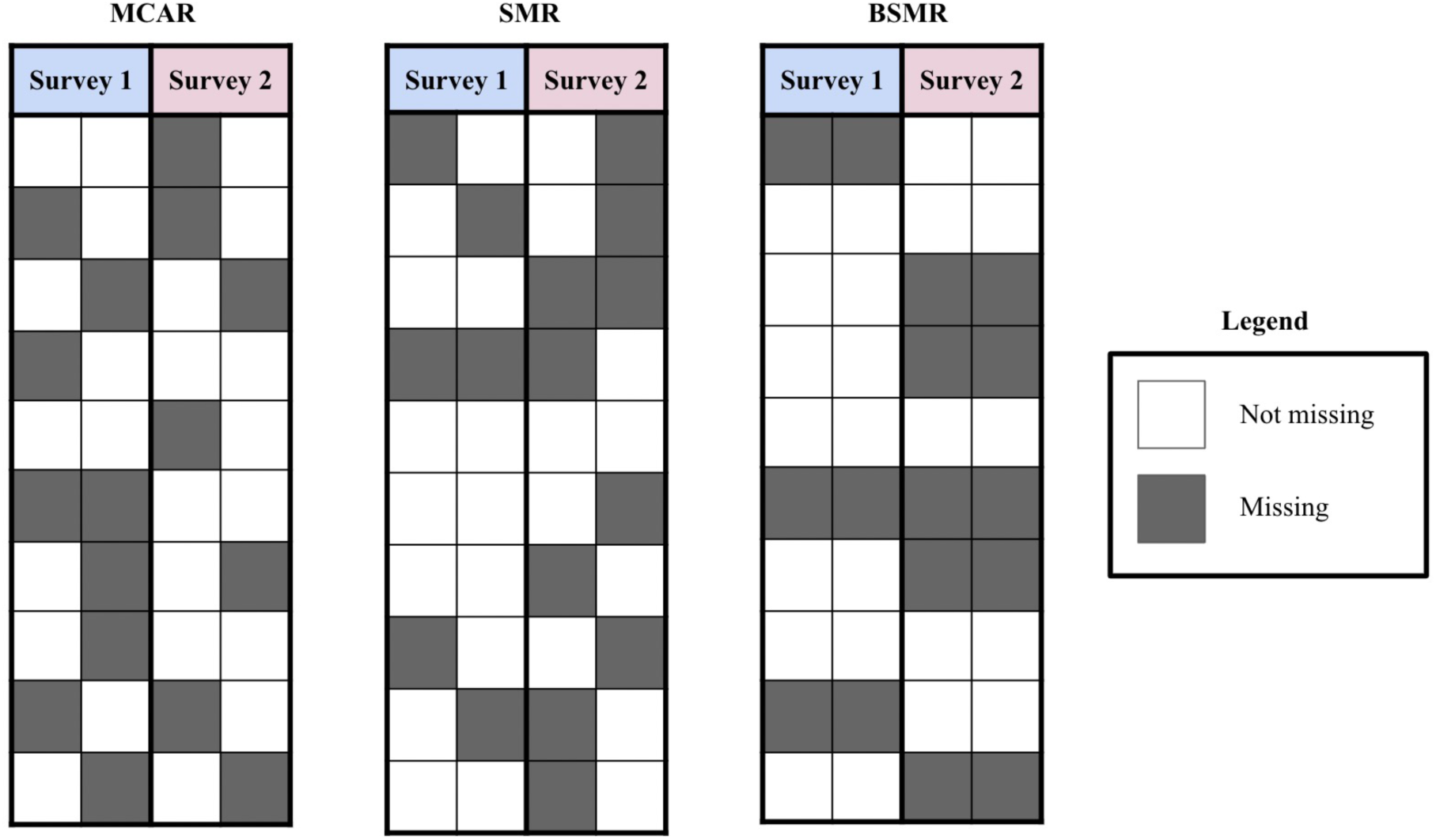
Visualization of the three missing data simulation scenarios explored in this study. On the left is Missing Completely at Random (MCAR) with a 40% missing rate. In the middle is Survey-Specific Missing Rate (SMR) with a 30% missing rate for Survey 1 and 50% missing rate for Survey 2. On the right is Blockwise Survey-Specific Missing Rate (BSMR) with a 30% missing rate for Survey 1 and 50% missing rate for Survey 2.

#### Missing Completely at Random (MCAR)

The first missing data simulation scenario, referred to as MCAR, introduces missingness completely at random by converting a specific percentage of the preprocessed complete dataset to missing. To observe the imputation performance as the missing rate gradually increases, MCAR was implemented with missing rates from 10% to 90% in 10% intervals for all variables in the dataset.

#### Missing Not at Random (MNAR): Survey-Specific Missing Rate

The second missing data simulation scenario is SMR, in which the proportion of missing values in each column is dependent on the survey type that it belongs to. SMR is tailored to mirror the missing rates in the full SPARK dataset by reusing the same proportions of missing values for each survey (**Table 1**).

#### Missing Not at Random (MNAR): Blockwise Missingness with Survey-Specific Missing Rate

The last missing data simulation scenario, referred to as BSMR, incorporates blockwise missingness with survey-specific missing rates. Instead of randomly selecting a specific portion of each column to be converted to missing as in SMR, a proportion of participants are randomly selected to have completely missing values for all surveys of a particular survey type. In other words, every column of a specific survey type contains the same missing rows. This resembles real data more closely when subjects skip the entire survey.

### 3. Machine Learning Imputation

For each missing data simulation scenario described in the previous section, multiple machine learning models were used to impute the missing values. The generated incomplete datasets were passed through the following imputation algorithms to compute the predicted values. A separate set of 10 datasets with 20% randomly selected missing values was used to conduct hyperparameter tuning on each of these models.

#### MICE

This study used the MICE ^12^ (version 3.16.0) package in R which employs a multiple imputation model. It uses a concept called Fully Conditional Specification, in which each incomplete variable is imputed by a different model. It generates multiple imputed datasets that are averaged to retrieve the final imputed data. Since MICE employs a regression-based approach, hyperparameter tuning was not performed.

#### KNN

KNNImputer is a method in Python’s Scikit-learn package ^22^ (version 0.22) and was used to study the KNN algorithm. KNNImputer predicts each sample’s missing values by using the average value from the closest data points in the training set. Hyperparameter tuning was used to select the optimal value for the number of nearest neighbors used during imputation.

#### MissForest

MissForest ^15^ (version 1.5) is an R package which uses a Random Forest approach to impute missing values, building multiple decision trees to make predictions using the other remaining features. By averaging several classification or regression trees, MissForest employs out-of-bag error estimates and can capture complex, non-linear relationships. Hyperparameter tuning was used to select the optimal values for the number of trees and the maximum number of iterations.

#### MIDAS

MIDASpy ^23^ (version 1.3.1) is a Python package that was used to study the MIDAS algorithm. It introduces additional missing values into a given dataset and restores these values using an unsupervised neural network called a denoising autoencoder. Then, the resulting model is used to predict the values of the original missing data. Similar to MICE, MIDASpy generates multiple imputed datasets that are averaged to retrieve the final imputed data. Hyperparameter tuning was used to select the optimal values for the input drop, layer structure, and number of epochs.

### 4. Evaluation of imputation performance

For each missing data simulation scenario, we introduced missingness into the complete dataset 10 different times as 10 separate trials. The values in **Table 1** correspond to the percentage of subject IDs in the full dataset (with missing values among participants with autism) who are not present in each specific survey. These missing rates were used when generating the missing datasets for the SMR and BSMR simulation scenarios.

The four models were used to impute the missing data, and these imputed values were compared with the true values in the preprocessed complete dataset. In each imputation trial, the RMSE values were calculated for each column using the `postResample` method from the `caret` package (Version 6.0-94) in R. The means of the RMSEs across all columns were aggregated to retrieve an overall RMSE. Then, these means were averaged across the 10 trials for each simulation setting. This resulted in a mean overall RMSE for each simulation scenario. These error values were then compared for every simulation scenario between each imputation method.

SCQ summary score, RBS-R summary score, and DCDQ summary score evaluate the social communication function, severity of repetitive behaviors, and motor functions respectively in study participants with autism. They were calculated based on corresponding questionnaires. The RMSE values of these specific mental and behavior summary scores were also compared between the four imputation methods across each simulation scenario.

Lastly, the total computation time was assessed for the four imputation methods during the BSMR simulation scenario, which was chosen since it is closest in nature to missingness in real survey data.

## Results

### 1. Overview of full dataset and missingness patterns

The full dataset used in this study consists of 117,099 study participants with autism. 51.3% of the participants did not complete SCQ survey which screens for social functioning, 63.8% did not complete RBS-R survey on repetitive behaviors, and 72.9% did not complete DCDQ survey on motor functions (**Table 1**). 34,067 participants have medium missing rates between 20% and 80% among 363 total questions (**Table 2**). 37,710 participants exhibit low missing rates (<20%) whereas 45,322 participants exhibit high missing rates (>80%, **Table 2**).

**Table 2.** Sample characteristics by low (<20%), medium (20%-80%), and high (>80%) missing rate in SPARK. Proportion of missing variables for each subject was calculated in the full dataset of this study containing 117,099 total participants with autism. Organized by different demographics including sex, age, and race.

|  | Missing Rate |  |  |  |
| --- | --- | --- | --- | --- |
|  | Low missing rate<br>(<20%) | Medium missing rate<br>(20-80%) | High missing rate<br>(>80%) | p-value |
| **Number of Subjects** | 37710 (32.2) | 34067 (29.1) | 45322 (38.7) |  |
| **Sex (%)** |  |  |  | <0.001 |
| Male | 29460 (33.5) | 24030 (27.3) | 34412 (39.1) |  |
| Female | 8250 (28.3) | 10037 (34.4) | 10910 (37.4) |  |
| **Age (%)** |  |  |  | <0.001 |
| <2 years | 456 (28.5) | 636 (39.7) | 509 (31.8) |  |
| 2-5 years | 9773 (38.0) | 6189 (24.1) | 9726 (37.9) |  |
| 6-11 years | 16511 (39.1) | 9230 (21.9) | 16463 (39.0) |  |
| 12-18 years | 10966 (38.4) | 6217 (21.7) | 11401 (39.9) |  |
| >18 years | 4 (~0.0) | 11795 (62.0) | 7223 (38.0) |  |
| **Race (%)** |  |  |  | <0.001 |
| White | 28727 (47.3) | 17968 (30.0) | 14093 (23.2) |  |
| African American | 2063 (37.8) | 1373 (25.2) | 2021 (37.0) |  |
| Asian | 876 (35.0) | 645 (25.7) | 988 (39.4) |  |
| Native American | 180 (37.4) | 141 (29.3) | 160 (33.3) |  |
| Native Hawaiian | 55 (43.0) | 29 (22.7) | 44 (34.4) |  |
| Multiple Races | 4155 (48.3) | 2203 (25.6) | 2249 (26.1) |  |
| Other | 1654 (4.2) | 11708 (30.0) | 25767 (65.9) |  |

When compared to female participants, there are slightly more male participants with high and low missing rates. Around 39% of male participants have high missing rates, which is slightly larger than the 37% of female participants. While 33.5% of male participants have low missing rates, only around 28% of female participants have low missing rates within this range.

For individuals between ages 2 and 18, around 22% of these participants have medium missing rates. The missing rates of these individuals are more concentrated towards extreme values, since around 39% have either low or high missing rates and 22% exhibit medium missing rates. For individuals below 2 years of age, around 40% have medium missing rates. Around 62% of individuals above 18 years of age have medium missing rates, whereas nearly 0% exhibit low missing rates.

Close to half of the self-reported White participants, Native Hawaiian participants, and individuals who identified as “Multiple Races” have low missing rates. The rates of missingness for self-reported African American, Asian, and Native American individuals are concentrated toward the extreme values, with more than 30% exhibiting high missing rates while less than 25% of the participants who were self-identified as White or “Multiple Races” reported high missing rates. Those who self-reported themselves as an “Other” race exhibit large amounts of missingness, since around 66% have missing rates larger than 80%.

### 2. Sample Characteristics of Complete Dataset and Simulation of Three Missingness Patterns

To assess the imputation performance of the four popular missing data imputation methods (MICE, KNN, MissForest and MIDAS), we first obtained a preprocessed complete dataset with 15,196 participants with autism (**Table 3**, details in Methods). Around 78% of participants with complete data are male and 22% are female. The male to female ratio is 3.5:1, which aligns with the sex ratio among subjects with autism in the general population. About half of the individuals with complete data are between 6-11 years of age. Only 0.4% of subjects are under 2 years of age while none are above 18. 79% of participants were self-identified as White. The category with the second largest number of participants is “Multiple Races” (10.9%), followed by African American (4.3%), followed by “Other” (3.5%), followed by Asian (2.2%). The number of participants who are Native American or Native Hawaiian are below 1%. In the preprocessed complete dataset, the SCQ, RBS-R, and DCDQ scores have average values of 21.72, 35.16, and 37.87 respectively.

**Table 3.**
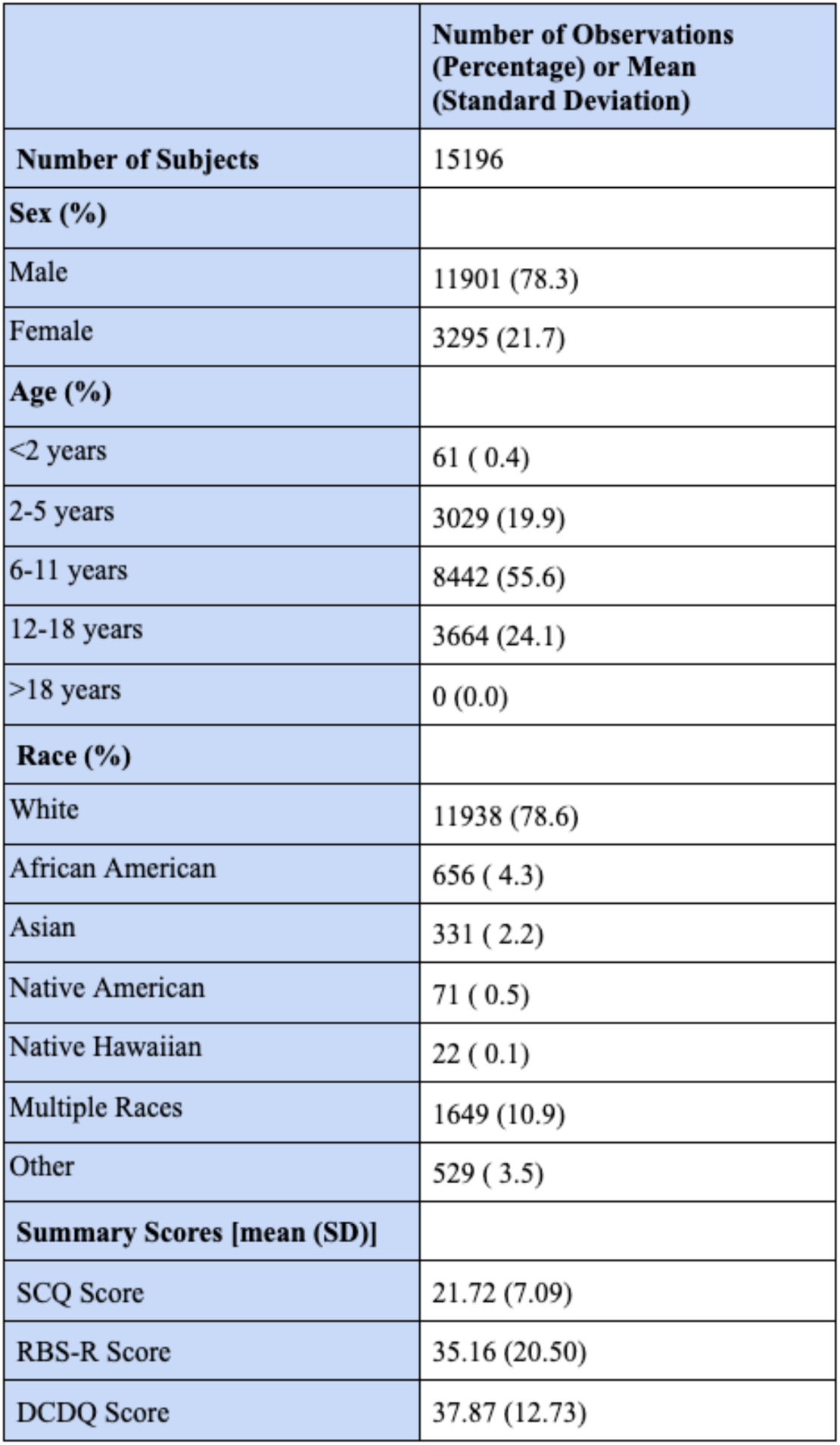
Sample characteristics in the preprocessed complete dataset containing 15,196 participants. This table includes the number of observations and percentage breakdowns of sex, age, and race as well as means and standard deviations of the Social Communication Questionnaire (SCQ), Repetitive Behavior Scale-Revised (RBS-R), and Developmental Coordination Disorder Questionnaire (DCDQ) summary scores.

To assess the performance of the missing data imputation methods, missing values were introduced to the preprocessed complete dataset with 15,196 participants with autism. First, to simulate the scenario on MCAR, a random subset of values across the entire dataset were converted to missing values. 10 incomplete datasets were generated for each missingness percentage (10%-90%). Second, to examine the performance of the imputation methods on MNAR patterns, 10 incomplete datasets were randomly generated for the SMR and BSMR simulation scenarios separately. When doing so, the missing rates in the original SPARK dataset were used (**Table 1**) to reflect the missingness distribution present in the real data.

### 2. Performance of Imputation on Overall Dataset

The four imputation methods were applied to the incomplete datasets in each of the three simulation scenarios (Figure 3). The imputed values were compared with the actual values in the complete dataset, and the RMSE values were calculated. Lower RMSE values correspond to higher accuracy in missing value imputation.

**Figure 3.**
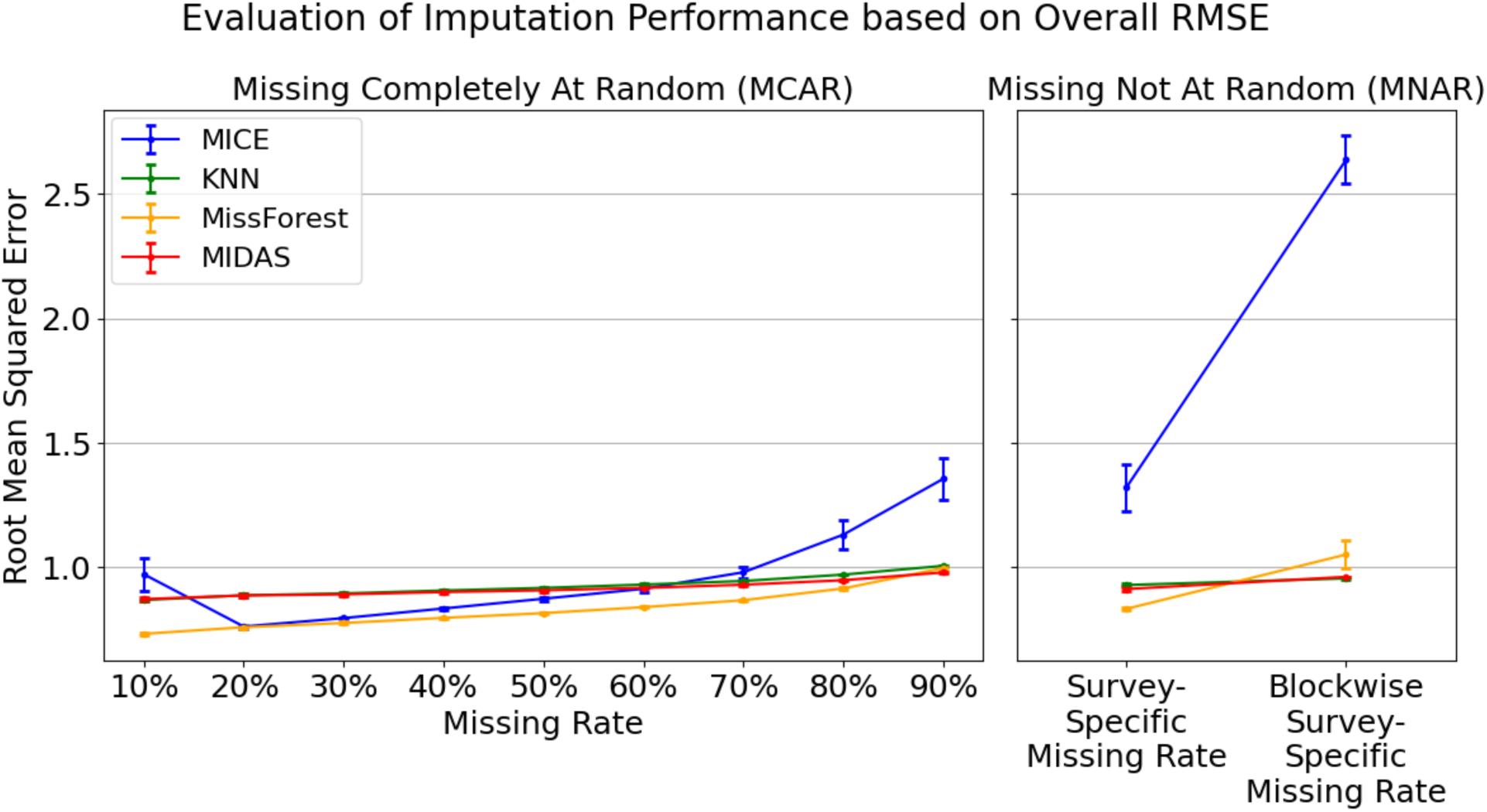
Evaluation of imputation performance based on overall RMSE. Values across the 10 trials using the MCAR simulation scenario (left). Overall RMSE values across the 10 MNAR trials in the Survey-Specific Missing Rate (SMR) and Blockwise Missingness with Survey-Specific Missing Rate (BSMR) simulation scenarios (right).

In the MCAR scenario, the imputation error for all models generally rose as the missing rate increased. MissForest has the lowest overall RMSE (ranging between 0.73 and 1.0), outperforming the other methods especially when missing rate was low (Figure 3, left panel). However, as the percentage of missing values increased, the performance of KNN and MIDAS became comparable to that of MissForest. MICE outperformed KNN and MIDAS between 20% to 60% of random missingness but performed considerably worse than all other models for the remaining missing rates.

In the MNAR scenarios, all models exhibited an increase in imputation error in the BSMR scenario when compared to SMR. MissForest produced the lowest error rate in the SMR scenario, with an RMSE of 0.83, but did not perform as well during the BSMR scenario that simulated blockwise missingness. MissForest also exhibited larger variations in RMSE (standard deviation = 0.056) in the BSMR scenario than in the SMR scenario (standard deviation = 0.0043). For the BSMR scenario, KNN and MIDAS performed the best with an average RMSE of 0.96. The variability of the RMSE was also relatively low for both methods, with a standard deviation of 0.0066 for KNN and 1e-6 for MIDAS. MICE performed worse than the other imputation methods in both SMR and BSMR scenarios. Especially in the BSMR scenario, the RMSE value was significantly higher at 2.64 with a relatively large standard deviation of 0.098.

For every simulation scenario, the difference in imputation performance on overall RMSE between KNN and MIDAS was marginal. Both models produced very similar results throughout the experiment and for each simulation scenario besides BSMR, they typically performed slightly worse than MissForest.

### 3. Performance of Imputation on Mental and Behavioral Summary Scores

For every simulation scenario, the mean and standard deviations of RMSE values for the SCQ, RBS-R, and DCDQ scores were computed across the ten trials as displayed in Figure 4. The relative performance of the four models was generally consistent across the three summary scores.

**Figure 4.**
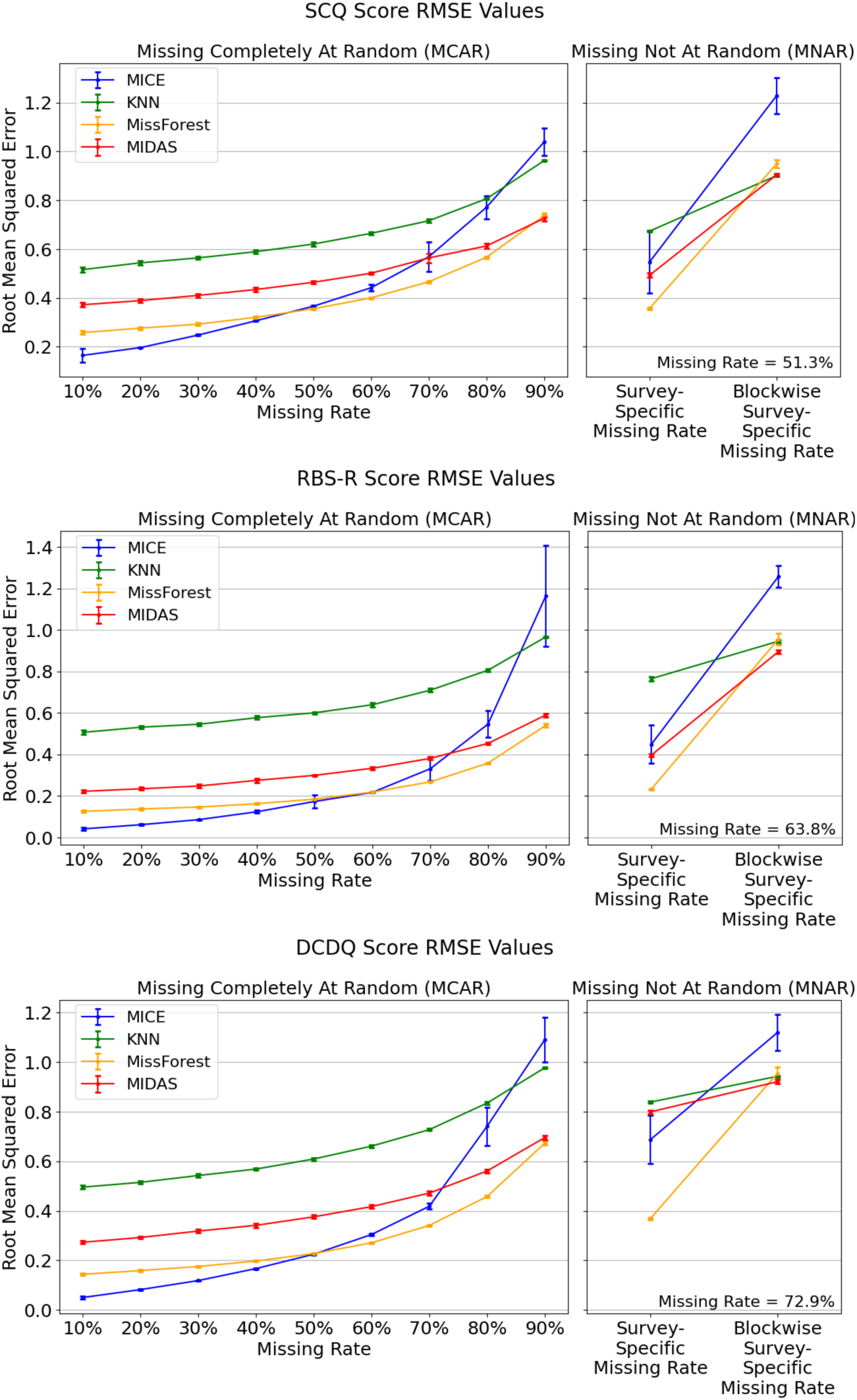
Imputation performance on summary scores from mental health surveys. Root Mean Squared Error (RMSE) values for imputing the Social Communication Questionnaire (SCQ) score across the MCAR and MNAR trials (top). RMSE values for the Repetitive Behavior Scale-Revised (RBS-R) score across the MCAR and MNAR trials (middle). RMSE values for the Developmental Coordination Disorder Questionnaire (DCDQ) score across the MCAR and MNAR trials (bottom).

In the MCAR scenario, MissForest consistently outperformed KNN and MIDAS when imputing all three summary scores. The MICE model exhibited a steep incline in error as the missing rate was incremented. It performed the best until the missing rate was increased to 50%, after which it was surpassed by the remaining models. MICE is ideal for lower rates of random missingness but begins to perform exponentially worse as the rate gets larger. In fact, the MICE model produced the largest RMSE among the four methods at a 90% missing rate. For missing rates that are 50% and above, MissForest is the ideal model since it had the lowest errors among the four methods.

The MissForest model performed the best in the SMR scenario. However, each method, especially MICE and MissForest, exhibited error rates that rose sharply when the missing values became blocked by survey type in the BSMR scenario. In the BSMR scenario, KNN and MIDAS exhibited the lowest error rates with MissForest performing slightly worse. MICE performed considerably worse than the remaining models in the BSMR scenario.

### 4. Computational Time

When comparing the computational times of the four models, the BSMR simulation scenario was used since this environment most closely resembles the missingness patterns in the real data when participants skip an entire survey in SPARK.

As shown in Figure 5, MIDAS and KNN not only had similar overall error rates, but also exhibited comparable imputation times of around 10 to 13 minutes. MissForest had a median imputation time of slightly less than 30 minutes. On the other hand, MICE had a median imputation time of around 285 minutes, which was significantly larger than those of the remaining models.

**Figure 5.**
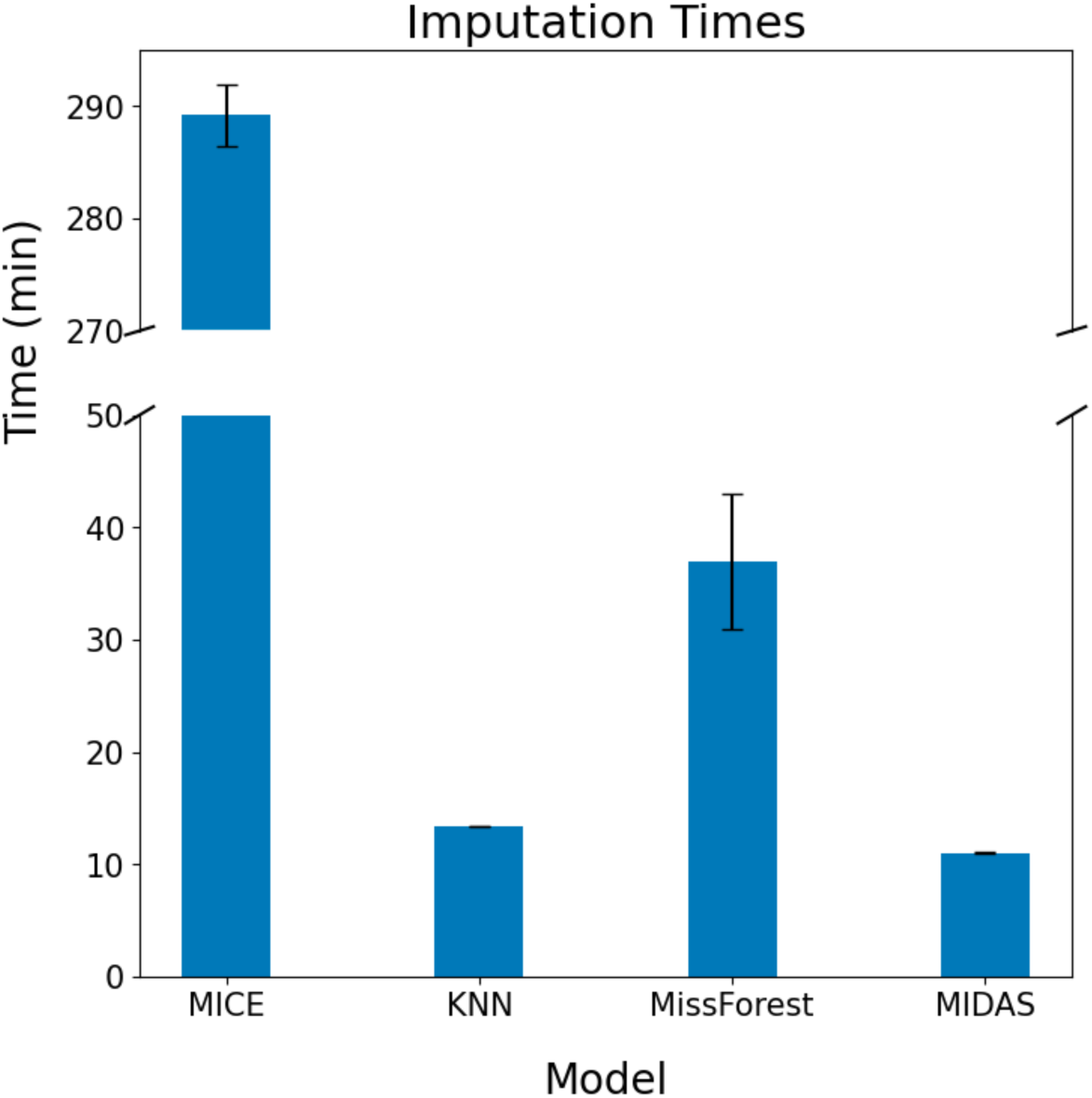
Total imputation times (in minutes) and standard deviations of each model for the 10 trials in the Blockwise Missingness with Survey-Specific Missing Rate (BSMR) scenario. Total sample size is 15,196.

## Discussion

The establishment of biobank databases has enabled the collection of self-reported mental and behavioral surveys at scale^1-3^. SPARK has gathered social and behavioral survey data from about 100,000 individuals^1^ and there is ongoing collection of more survey data on existing participants. UK Biobank has measurements on lifetime depressive disorder, cognitive function, attention, and impulsivity from about 150,000 participants ^2,24,25^. All of Us also has strategic plans to collect mental and behavioral surveys at scale^3^. However, the data quality and statistical power are compromised by missing data. Recent advances in machine learning methods have inspired novel missing data imputation approaches with increased accuracy and computational efficiency ^12-15^. Previous studies either have not reviewed these newly developed imputation methods or have not focused on assessing imputation accuracy in mental and behavioral surveys that exhibit blockwise missing structures ^17-21^.

Our study provided insights on the missingness pattern in SPARK, a large-scale cohort with autism, and assessed the imputation accuracy and computational time of four popular missing data imputation methods – MICE, KNN, MissForest and MIDAS. We did this by simulating three missingness scenarios in mental and behavioral surveys, including SCQ, RBS-R and DCDQ. We observed that 50%-70% of participants with autism did not complete SCQ, RBS-R and DCDQ surveys and the dataset exhibited blockwise missing structures. The missing rates also varied by sex, age, and race. Overall, KNN and MIDAS showed relatively stable performance with increasing missing rate in the MCAR scenario and slightly higher imputation error when blockwise missingness is introduced in the MNAR scenarios. The error rate increased more significantly in MICE and MissForest in both MCAR and MNAR scenarios, with a particularly notable surge in error rate for MICE when blockwise missing structures were introduced. When imputing SCQ, RBS-R and DCDQ summary scores in the MCAR scenario, MICE had the lowest error rate when the missing rate was low, while MissForest had the lowest error rate when the missing rate was high. However, in the presence of blockwise missingness in the MNAR scenario, MIDAS was consistently the best performing model across all three summary scores, with KNN and MissForest having similar or slightly higher error rates. Our results suggested that some models like MICE are sensitive to high missing rates and blockwise missing structures, while MIDAS and KNN may perform better in the overall dataset and specific summary scores in the presence of blockwise missingness. The average computational times for MIDAS and KNN to impute 15,196 subjects with blockwise missingness were about 10 minutes, about 35 minutes for MissForest, and about 290 minutes for MICE. These results highlight the computational efficiency in machine learning imputation algorithms even in highly complex neural network models in MIDAS. Newly developed imputation models have better optimization in their algorithms and take advantage of parallel computing to reduce the computational time.

Our results show the potential to impute missing data in large-scale databases with mental and behavioral surveys, especially imputing summary scores based on medical history and neurodevelopmental measures. When the data exhibits blockwise missingness, the imputation error increases but models such as MIDAS and KNN can still provide imputed results that are relatively stable and accurate. This shows that when a block of correlated variables in one survey is completely missing, other related surveys or medical history can also provide relevant information for imputation. The choice of imputation methods may depend on the overall missing rate and missingness patterns in a dataset.

The strength of our study is that we utilize a large-scale collection of mental and behavioral surveys in SPARK to simulate the missingness patterns, particularly with blockwise missing structures that are commonly observed in mental health databases. We also systematically assessed the latest missing data imputation approaches like MIDAS. Our limitation is that the complete data with missing data simulation primarily comes from adolescents. Despite the inclusion of various racial groups in the simulation, most participants are white. Assessment in other types of large-scale mental and behavioral surveys with adults and minority groups is warranted for future studies.

Missing data imputation is widely used in national surveys with mental and behavioral surveys. For example, the National Survey on Drug Use and Health (NSDUH) has been providing imputation-revised variables by the predictive mean neighborhood methods since 1999^26^. There is also the recent phenotype imputation model developed in the UK Biobank, which has shown increased power for genetic studies ^27^. As biobanks and national surveys collect more large-scale data on mental and behavioral surveys, missing data imputation will produce more accurate imputed values and become an integral part of analysis to maximize the use of the data.

Our study underscores the efficacy of advanced imputation techniques, such as MIDAS and KNN, in addressing missing data within large-scale mental and behavioral surveys. Our findings showcase that for similar databases with mental and behavioral surveys on autism, dementia and other disorders, machine learning-based imputation methods can be leveraged to effectively recover missing information. This study demonstrates that machine learning methods offer increased performance and faster computation times over traditional algorithms. The performance of these advanced imputation techniques demonstrates their potential to optimize analyses and advance research in mental and behavioral disorders.

## Data availability

SPARK Phenotype Dataset is accessible through application at SFARI Base (https://base.sfari.org)

## Code availability

All software used in this study is publicly available. The code for simulations and analysis can be found at https://github.com/AprilShuLab/MissingDataImputation.

## Acknowledgements

We are extremely grateful to the thousands of individuals and families who are participating in the SPARK. We thank the sites, staff and volunteers of the SPARK Clinical Site Network and SFARI for their invaluable contributions.

## Author contributions

Preethi Prakash conducted the entire analysis and wrote the manuscript and Dr. Chang Shu supervised this work. Dr. Kelly Street, Dr. Shrikanth Narayanan and Dr. Yufeng Shen provided guidance on the methodology, and Dr. Bridget Fernandez offered insights on clinical relevance.

